# Systemic inflammation and its associations in acute moderate-severe Traumatic Brain Injury: a cross-sectional study

**DOI:** 10.1101/2025.10.27.25338886

**Authors:** Lucia M. Li, Eleftheria Kodosaki, Chloe Xu, Amanda Heslegrave, Henrik Zetterberg, Neil Graham, Karl Zimmerman, Elena Garbero, Federico Moro, Sandra Magnoni, Guido Bertolini, David J. Loane, David J Sharp

## Abstract

Traumatic Brain Injury (TBI) triggers an acute systemic inflammatory response, which may contribute to poor long-term outcomes. Additionally, pre-existing factors associated with increased inflammation, such as age, may interact with this acute post-TBI inflammation to influence outcomes. Previous investigations of post-TBI inflammation have typically assessed small numbers of cytokines, but novel high-dimensional proteomic approaches can sensitively detect a broad range of inflammatory markers and more fully characterise post-TBI inflammation.

We analysed plasma from 84 participants in the BIO-AX-TBI cohort [*n*=37 acute, moderate– severe TBI (Mayo Criteria), *n*=22 acute non-TBI trauma (NTT), *n*=28 non-injured controls (CON)] on the Alamar NULISA™ Inflammation panel, assessing >200 inflammatory markers. The NTT group allowed differentiation of TBI-specific from general injury-related acute inflammatory responses. Inflammatory markers were correlated with plasma levels of NFL (neurofilament light), GFAP, total tau, UCH-L1 (all Simoa®) and S100B (Millipore); and subacute (10 days to 6 weeks post-injury) 3T MRI measures of lesion volume and white matter injury (fractional anisotropy).

Differential expression analysis identified 4 markers showing TBI-specific elevations in plasma levels (VSNL1, IL1RN/IL-1Ra, GFAP, IKBKG), whilst derangements in other inflammatory markers likely reflected a non-specific injury response. Higher VSNL1 levels were associated with greater lesion volume (r_s_=0.53), and higher IL1RN/IL-1Ra levels were associated with more white matter injury (r_s_=-0.66, both FDR-adjusted p<0.05).

The non-specific injury response was associated with functional outcome at 6-months – higher IL33 levels in those with good (Glasgow Outcome Scale-Extended, GOS-E, 5-8) versus poor (GOS-E 1-4) outcomes (W=47, FDR-corrected p=0.0024). To assess age-related effects, we calculated “inflammation age” by applying an Elastic Net model trained on a public healthy control dataset. The “age gap” (“inflammation age” minus calendar age) was greater in TBI than CON, and also greater in young participants.

In summary, the acute post-TBI inflammatory response is comprised of both TBI-specific and non-specific injury components. These inflammatory responses are associated with structural brain injury measures and overall functional outcome. We additionally find that age influences the acute inflammatory response. Our study highlights VSNL1, IL1RN/IL-1Ra and IL33 as potential inflammatory mediators of post-TBI pathophysiology.

## INTRODUCTION

Traumatic brain injury (TBI) causes long-lasting problems affecting not only the brain, but also seems to trigger systemic processes, such as inflammation. A peripheral pro-inflammatory response (e.g. increased plasma levels IL6, TNF, IFNy) is consistently reported in acute TBI, with higher levels associated with worse neurobehavioural and global outcomes^1–8^. Aberrant systemic inflammation is seen in many systemic and neurobehavioural health problems, e.g. depression^9,10^, and cross-talk between central and peripheral immune systems is implicated in neurodegenerative disease^11^. Therefore, post-TBI systemic inflammation may contribute to the development of multisystem problems.

To date, small numbers of inflammatory markers or pathways have been explored in previous clinical studies, and the TBI-specificity of the inflammatory response is often not explicitly assessed. One previous study used mass spectroscopy to identify >1000 proteins that had altered levels in the CSF of acute TBI patients, compared to non-injured controls^12^. This study highlights the potential for proteomic approaches to identify previously unknown inflammatory pathways impacted by TBI. High-dimensional multiplex immunoassay panels enable simultaneous detection of a wide range of targets, typically with higher sensitivity, lower cost and with simpler workflows than mass spectroscopy approaches. This makes them particularly suitable for discovery work exploring the full range of disease pathophysiology and identify potential intervention targets, in a cost-effective way. We have previously used the Alamar NULISA™ CNS diseases panel to identify acute, TBI-specific derangements in plasma levels of many proteins not previously seen in human TBI^13^. However, only a limited number of inflammation markers are included on the CNS diseases panel.

Here, we tested acutely-collected plasma samples from a sub-group of the BIO-AX-TBI study^14^, a longitudinal multi-modal TBI cohort study, using the Alamar NULISA™ Inflammation panel for the first time in acute TBI. This panel assess 250 inflammatory markers, enabling a more comprehensive characterisation of the acute peripheral inflammatory response to TBI than has been previously possible. Including both non-injured (CON) and non-TBI trauma (NTT) controls also enabled us to explicitly investigate the TBI-specificity of the inflammation response. We additionally investigated which inflammatory markers were associated with neuroimaging measures of intracranial injury and clinical outcome. Finally, we derived a composite measure of overall inflammatory response, the “Inflammation Age”, as a way to investigate the impact of age on post-TBI inflammation.

We hypothesised that the acute systemic inflammatory response post-TBI would comprise elevations in pro-inflammatory cytokines and chemokines. We predicted that this response would comprise both TBI-specific and non-specific injury components. We additionally hypothesised that TBI-specific inflammatory markers will be associated with MRI measures of intracranial injury and functional outcomes. Finally, that “Inflammation Age” is increased after TBI, indicative of an acute inflammatory response to injury, but that this increase is influenced by age.

## METHODS

### Participant Cohort

Plasma samples (n=84) were available from a subgroup of BIO-AX-TBI study^14^, which prospectively enrolled individuals aged 18–80 years with moderate–severe TBI (Mayo Criteria^15^) following hospital admission, together with a non-TBI trauma (NTT) comparison group and healthy non-injured controls (CON) (TABLE1). Further inclusion/exclusion criteria and injury mechanisms are detailed in Supplementary Information. Eleven TBI patients had pre-injury comorbidities, including hypertension, non-insulin dependent Type II diabetes, arrhythmia, and prior alcohol misuse (full comorbidity list, see SI Table 1).

**TABLE 1:**
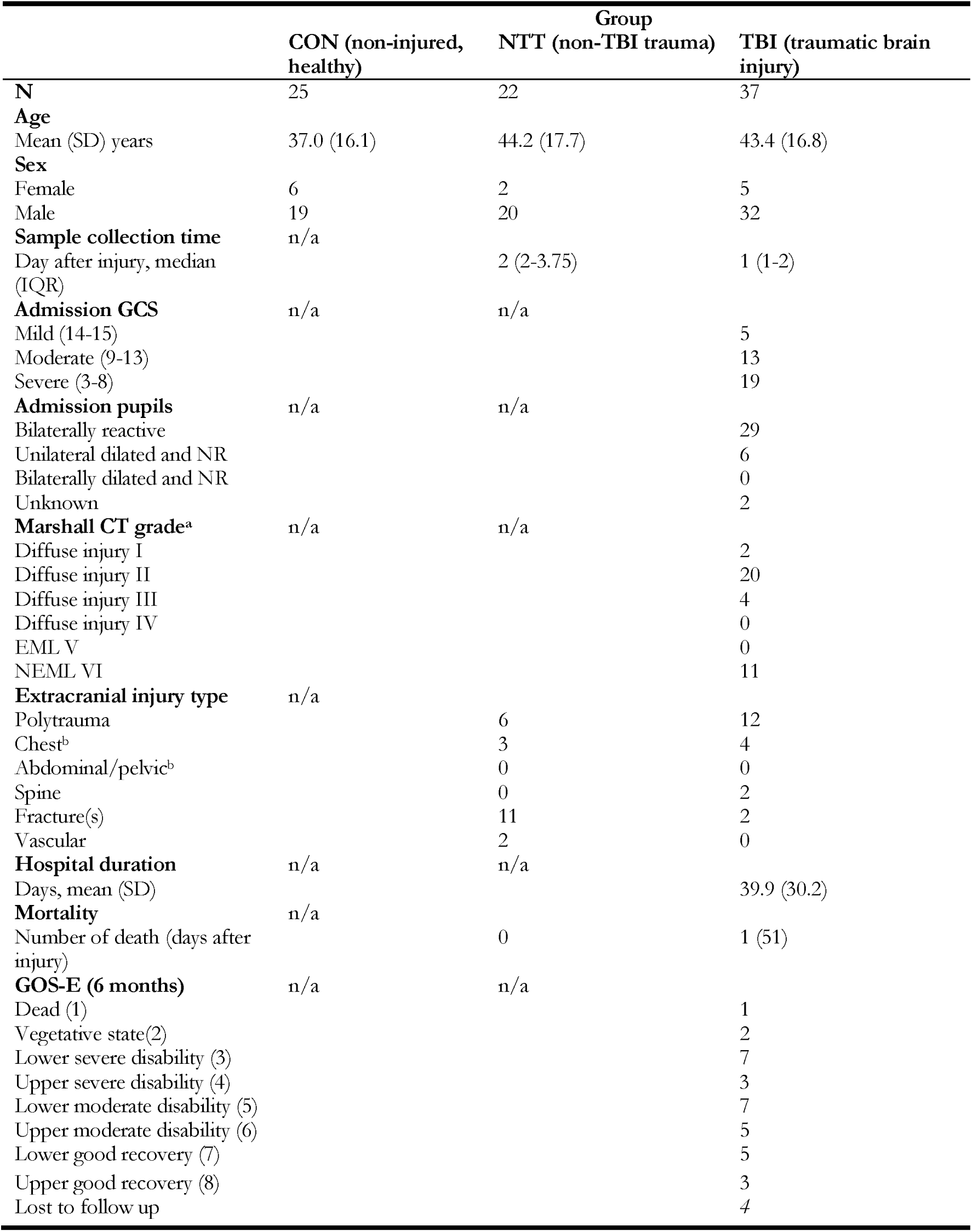
Clinical and demographic information. EML=evacuated mass lesion; NEML=no evacuated mass lesion; GOS-E=Glasgow Outcome Scale Extended. ^a^On initial CT. ^b^Indicates injury of chest or abdominal or pelvic organs; isolated rib or pelvic fractures with no underlying organ damage was classified under Fractures.

### MRI Acquisition and Analysis

N=34 TBI (29M:5F, mean age=43.1years, sd age=17.1) and all CON participants had subacute 3T MRI scans (10 days to 6 weeks post-TBI, to allow for the practicalities of acquiring MRI in acutely unwell patients). We included lesion volumes and measures of white matter injury in this analysis. MRI was acquired, preprocessed and analysed for the BIO-AX-TBI study (protocol details^14,16^, full details *Supplementary Information*). Diffusion tensor imaging was available for 28 TBI (23M:5F, mean age=42.5years, sd=17.1) and 23 CON participants (19M:4F, mean age=36.2years, sd=16.2). Two TBI patients could not have a subacute MRI due to practical difficulties, 1 patient did not tolerate the scan. Of those who had DWI, diffusion data for 6 TBI and 2 CON were not usable due to poor data quality or artefacts. White matter injury was assessed with *z*-scored mean fractional anisotropy (FA) across the whole skeletonised white matter after registration of diffusion scans to DTITK space^17^, and using a tract-based spatial statistics (TBSS) approach to generate voxelwise maps of FA. Z-scores were calculated by comparing values from patients and controls scanned on the same scanner. Mean *z*-scored FA was extracted for the corpus callosum and whole white matter skeleton.

### Blood sample processing

Plasma samples were collected, processed and frozen within 10 days post-injury^7^, and analysed on the Alamar Biosciences’ NULISA™ Inflammation Panel^9^ in one batch. They were also analysed on the OLINK® Target Inflammation 96 and Alamar NULISA™ CNS Diseases panels, as previously described^13^. All samples were immediately taken to labs for processing and storage at −80°C until analysis. To ensure roughly balanced group sizes, all available NTT samples were included, while TBI and CON samples were randomly selected after excluding GFAP outliers (more than 3 standard deviations outside mean for that cohort) (*Supplementary Information*)^13^.

The NULISA™ Inflammation panel offers high-dimensionality, enabling testing of a wide range of inflammatory markers with one low-volume sample, while the OLINK® panel provided cross-platform validation. NULISA™ and OLINK® profiling in these samples has been described in detail previously^13^. In brief, samples tested on the Alamar NULISA™ Inflammation panel were tested after 2 previous freeze-thaw cycles. Normalisation takes place against both internal and inter-plate controls. Biomarker data for both Alamar and OLINK® assays are reported in relative protein concentrations, given as normalised protein expression (NPX) values for the OLINK® panel, and NULISA Protein Quantification (NPQ) units for the Alamar panel. Both are arbitrary units on a log2 scale, with the data being normalised to minimise variation.

### Statistical analysis

All statistical analyses were performed in R (version 4.3.2), using RStudio (2021).

We carried out Differential Expression (DE) analysis on the Alamar panel data to identify proteins whose plasma levels are affected by TBI. DE is a statistical approach to detect and identify, on a wide scale, biological markers whose expression varies between different groups. We used the *limma* package from *Bioconductor*, using the linear model option. For the core DE analysis, we included age and sex as covariates. Although average sample collection time was not significantly different between TBI and NTT, given the potential for inflammatory cytokines to change over the first few days post-TBI^18–20^, we specifically investigated the potential impact of sample time collection with an additional DE analysis, between TBI and CON cohorts only, adding sample day as a third covariate. FDR correction (the Benjamini-Hochberg method) was done for multiple comparisons across all proteins assayed. Proteins with ≥2-fold change and adjusted p<0.05 were considered significantly differentially expressed between each group pair.

Spearman correlations assessed relationships between proteins identified by the core DE analysis as having TBI-specificity with each other, and with MRI measures of injury (lesion volumes, whole-skeleton and corpus callosal fractional anisotropy (FA) z-score and corpus callosum FA z-score), within the TBI group only. P-values from correlation analyses were FDR-corrected for multiple comparisons within the MRI analysis and protein-protein correlation analysis separately. We used the Wilcoxon rank sum test to investigate differences in FA between TBI and CON cohorts (as TBI FA is non-normally distributed in our cohort).

To investigate associations between acute inflammation post-TBI and global outcomes, we compared acute levels of proteins identified by the core DE analysis as having TBI-specificity between TBI participants with GOOD (Glasgow Outcome Scale Extended^21^ [GOS-E] score=5-8) versus POOR (GOS-E=1-4) outcomes at 6 months (available in 33 TBI participants, 28M:5F, mean age=43.8years, sd age=17.3). We used Mann-Whitney U tests, FDR correction for multiple comparisons.

In an exploratory analysis, calculated “Inflammatory Age” for each participant as a measure of overall inflammatory status. We used Elastic Net (*cv.glmnet,* alpha=0.1) to train a predictive model that identified inflammatory markers associated with age, using a normative dataset comprising 485 individuals aged 18-89 years^22^. We then applied the coefficients and intercept to study participants to derive their “Inflammation Age” and “age gap” (“Inflammation Age” minus calendar age). A linear model (*lm*) investigated whether “age gap” was different between groups, with calendar age, sex, and interaction between group and calendar age as covariates. We used Spearman correlations to investigate the relationship between “Inflammation Age” and acute plasma levels of neuronal/astroglial markers, MRI measures, and 6-month GOS-E (FDR-corrected for multiple comparisons within neuronal/astroglial markers and MRI measures).

### Cross-validation

Pearson correlation analyses assessed correlations between markers assessed from different assay platforms (Alamar NULISAseq™ Inflammation Panel against OLINK Target 96 Inflammation panel, and against Alamar NULISAseq™ CNS Diseases Panel).

## RESULTS

### Differential expression analysis reveals proteins involved in calcium regulation, IL1 and NF-kB pathways

Core differential expression (DE) analysis identified 12 inflammatory markers whose plasma levels changes showed TBI specificity (FIG2A, 2B). Six markers had significantly higher plasma levels in acute TBI, compared to both acute NTT and CON: astroglial marker GFAP, cytokine receptor antagonist IL1RN, calcium sensor VSNL1, neuropeptide CALCA, and inflammatory/immune regulators TIMP1 and IKBKG. Six proteins had significantly elevated plasma levels in both acute TBI and NTT compared to controls, but with greater elevation in TBI compared to NTT: cytokines and growth factors IL6, LIF, PTX3, and IL33, its receptor IL1RL1, and immune regulator CHI3L1. Volcano plots demonstrate how plasma expression of proteins differed between each pair of groups (FIG1A), with the Venn diagram summarising proteins that were differentially expressed between TBI and CON only, and also between TBI and NTT (FIG1B).

After additional adjustment for sampling day, GFAP, IL1RN, VSNL1 and IKBKG were significantly higher in acute TBI compared to NTT (FIG2C). These 4 proteins (highlighted in red in FIG1B) are therefore the markers with the most robust TBI-specificity. Full statistics are reported in *Supplementary Information*.

**Figure 1.**
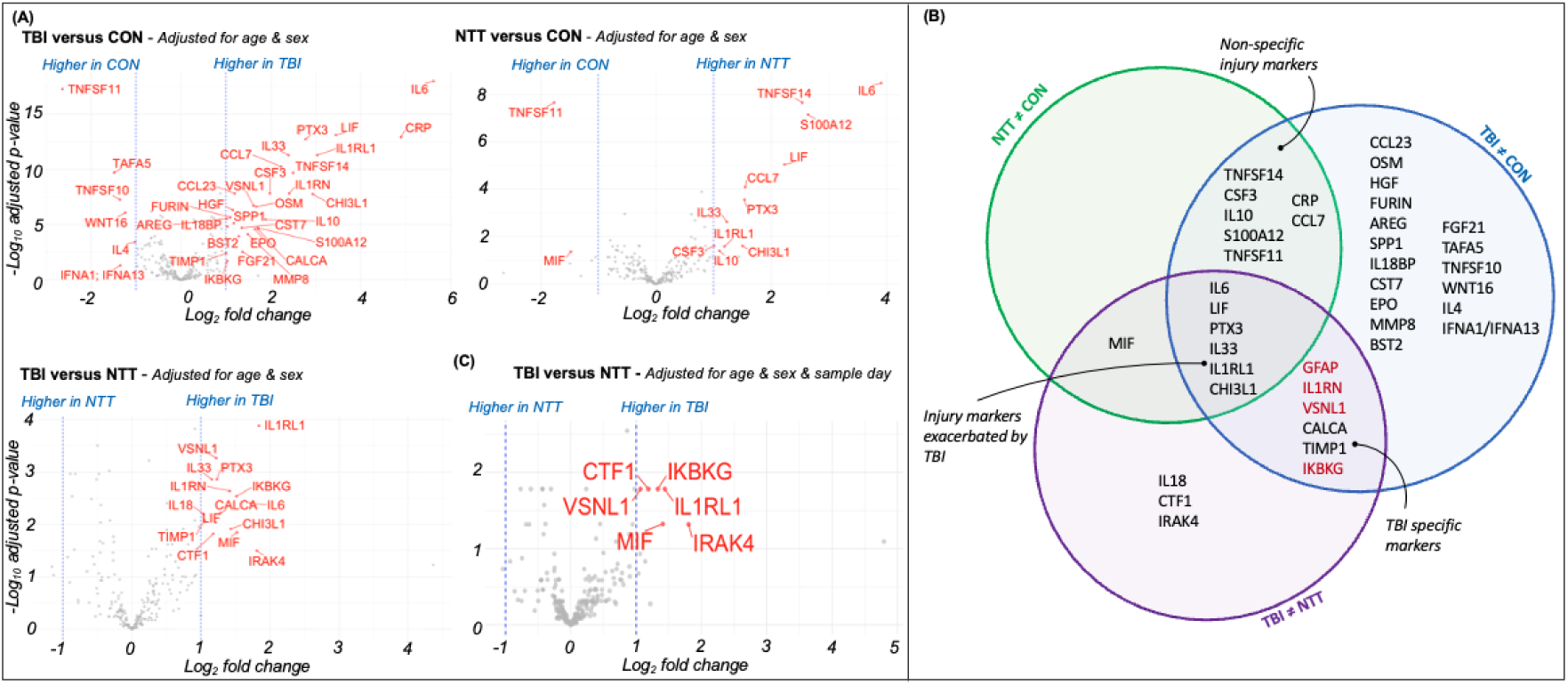
Differential expression analysis identifies TBI-specific changes in plasma inflammatory marker expression. Differential expression analysis visualising differences in plasma levels of inflammatory proteins between non-injured controls (CON), non-TBI trauma controls (NTT) and TBI. (A) Volcano plots showing significant differences in plasma protein expression between each pair of groups. The core differential expression analysis adjusted only for age and sex (top plus bottom left), and an additional differential expression analysis was performed (for TBI v NTT) adjusting for age, sex and sample day. Red dots denote significant proteins (defined as a log2 fold change≥|1| i.e. >2x difference between the two groups, denote by vertical dashed lines), with FDR-adjusted p<0.05. For visualisation clarity, GFAP is not represented on the volcano plot as it has a much higher fold change between TBI v CON and TBI v NTT than the other proteins. GFAP levels for each group are visualised in Supplementary Information Figure 1. (B) Venn diagram summarising the categorisation of proteins with significant pairwise group differences from differential expression analyses. Proteins in red denote proteins which continue to show differences between TBI and NTT after adjusting for age, sex and sample day.

#### Correlations between post-TBI inflammatory markers

There were multiple positive correlations between inflammatory proteins deranged in acute TBI (FIG2). The most pronounced associations were observed between cytokines of the IL6 family (IL6 and LIF, r_s_=0.92, FDR corrected p<0.01), and between these cytokines to other inflammatory markers (IL6 with IL1RN r_s_=0.72, LIF with IL1RN r_s_=0.70, IL6 with IL33 r_s_=0.7, all adjusted p<0.01).

**Figure 2.**
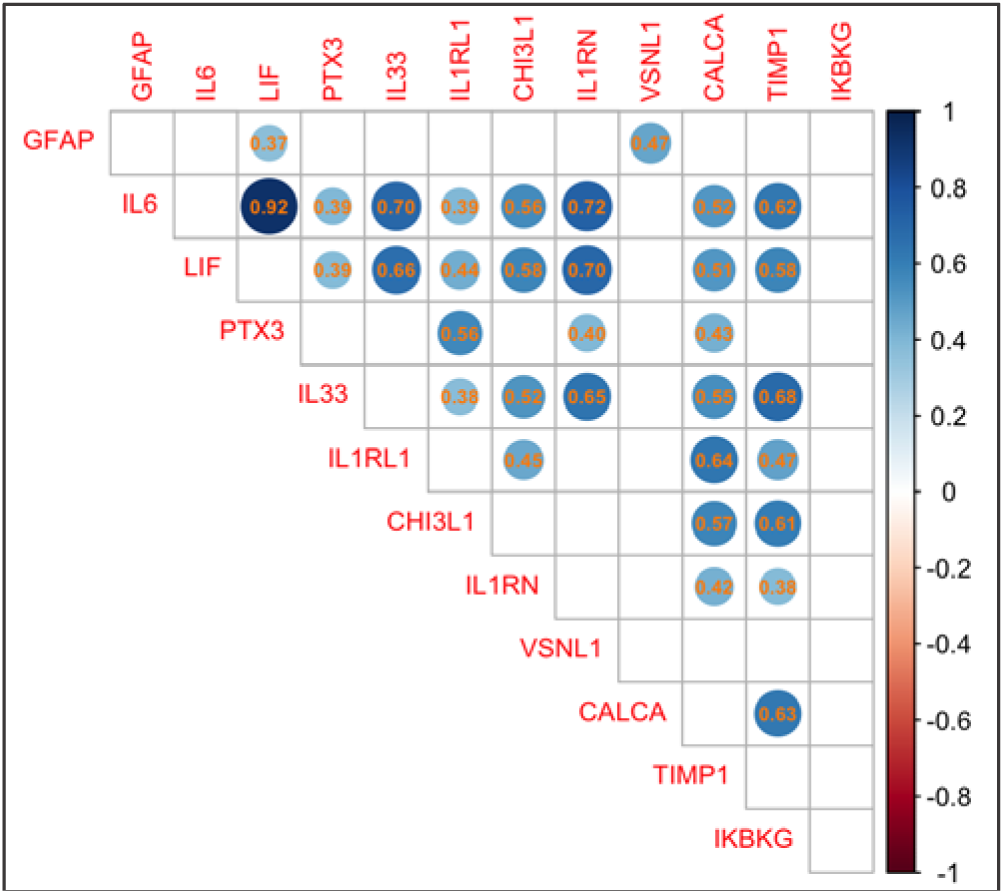
Correlations between TBI-specific proteins. Correlation matrix displaying relationships between TBI-specific proteins (identified by core DE analysis) within the TBI cohort. Spearman’s correlation coefficients are displayed within circles (larger and darker blue circle indicates stronger positive correlation). Only statistically significant correlations (FDR-adjusted p<0.05) are shown.

#### VSNL1 and IL1-receptor antagonist levels showed correlated with subacute MRI measures of injury

We then investigated whether inflammatory markers identified in the core DE analysis were associated with MRI measures of injury. White matter damage after TBI was assessed by reductions in fractional anisotropy (FA) from diffusion tensor imaging acquired in the subacute period (10 days-6 weeks post-injury). TBI patients had significantly lower mean FA values across the whole white matter skeleton (W=603, p<0.0001) and corpus callosum (W=590, p<0.0001) compared to CON, indicating white matter injury (FIG3A). Concentrations of VSNL1, a neuronal calcium sensor, were significantly positively correlated with subacute focal lesion volume (r_s_=0.53, p=0.024) (FIG3B). IL1RN levels were inversely correlated with corpus callosum FA (r_s_=–0.66, p=0.0047) ie. higher acute levels were associated with more white matter injury (FIG3B). Motivated by this result, we investigated the relationship between plasma IL1b, for which IL1RN is a receptor antagonist, and corpus callosum FA, but there was no significant relationship (r_s_=–0.04, p>0.05).

**Figure 3.**
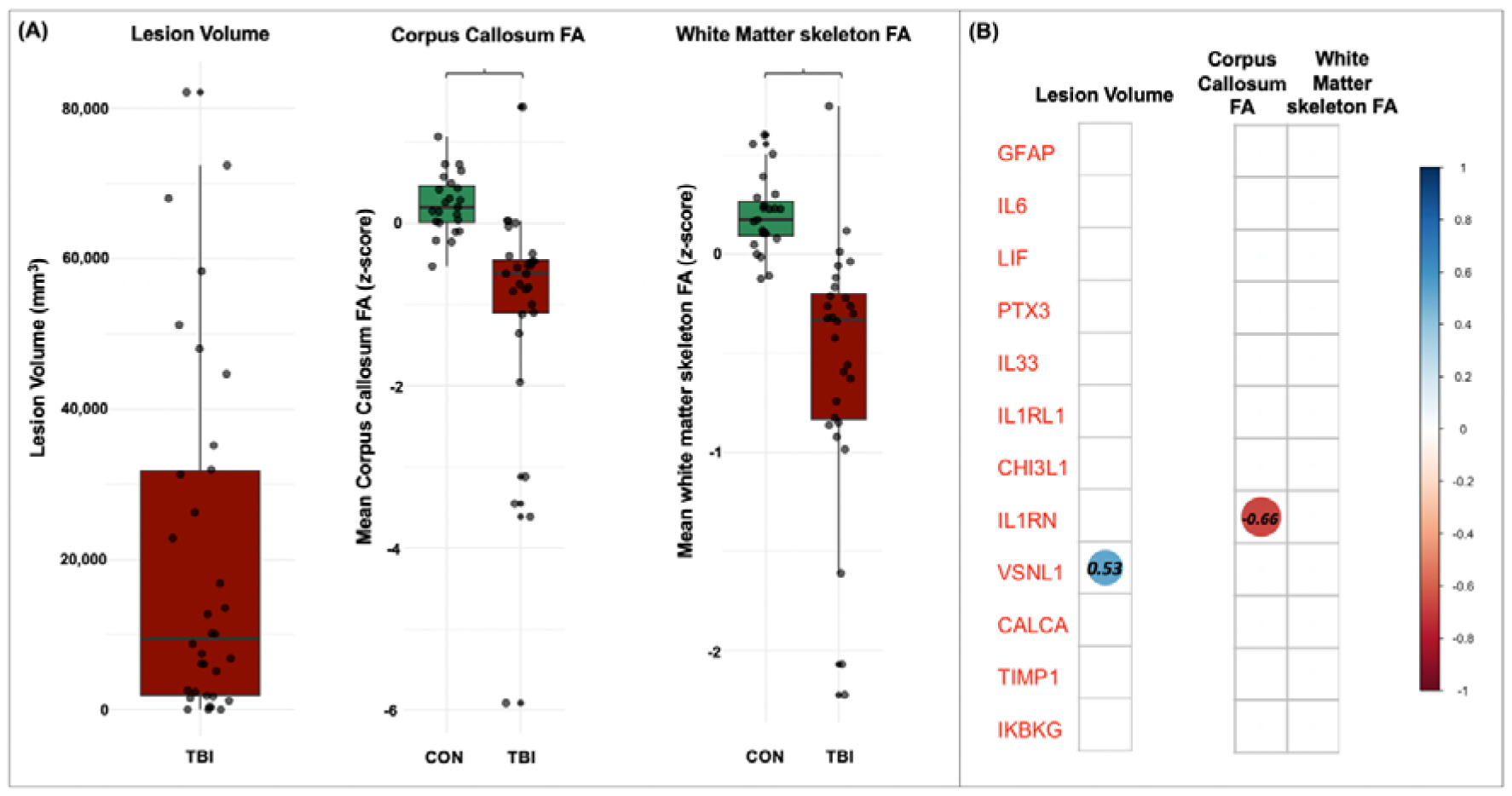
Relationship between TBI-specific proteins and neuroimaging features. (**A**) Lesion volumes in TBI, and white matter fractional anisotropy (FA) in non-injured healthy controls (CON) TBI participatns assessed on subacute MRI (10-42 days post-injury). *indicates p<0.0001 (Wilcoxon rank-sum test). Each dot represents a participant, box limits are the interquartile range (IQR), whisker limits are 1.5x IQR, midline is median. **(B)** Correlation matrices of Spearman correlations between TBI-specific proteins (identified by core differential expression analysis) and subacute MRI measurements in TBI patients: lesion volume (Lesion Vol, n=34), corpus callosum and whole white matter skeleton FA z-score (both n=28). Only statistically significant correlations (FDR-adjusted p<0.05) are shown.

#### Acute IL33 concentrations were higher in TBI patients with poor 6-month functional outcome

Acute IL33 plasma levels were significantly higher in TBI patients who had POOR functional outcome at 6 months, compared to those with good outcome (W=47, p=0.0024) (FIG4). To investigate the extent to which IL33 was independently associated with outcome, we performed a multivariate linear model that included age and clinical severity (the pre-hospital GCS), as covariates and confirmed that IL33 levels was significantly associated with GOS-E (*z*=2.406, p-0.016). Age and GCS were not. There were no associations between GOS-E and other proteins identified from the core DE analysis.

**Figure 4:**
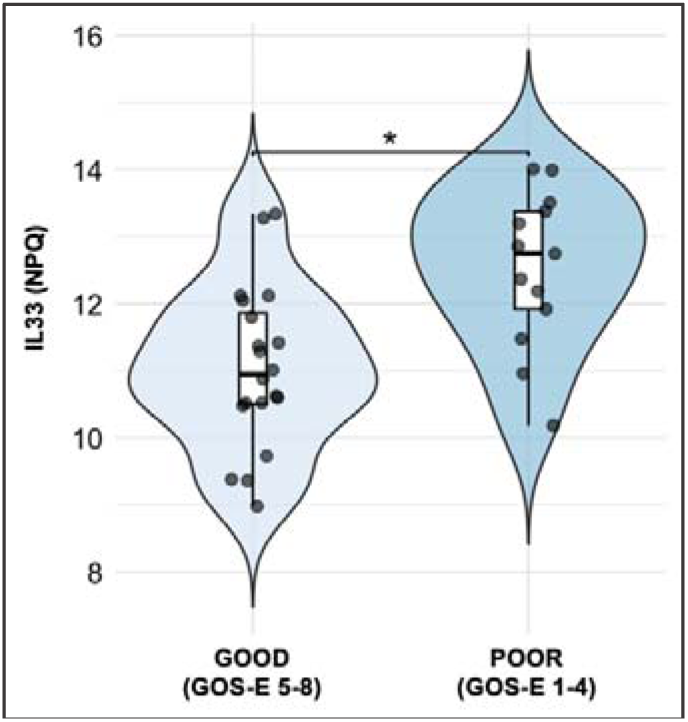
Acute plasma IL33 levels by 6-month outcome status. Acute plasma IL33 levels in TBI patients with POOR compared with GOOD 6-month Glasgow Outcome Scale-Extended (GOS-E) outcome category. Embedded boxplot limits denote interquartile range (IQR), midline denotes median, whiskers denote 1.5xIQR, each datapoint represents a participant. Statistical significance was determined using Mann-Whitney U test with FDR correction for multiple comparisons (*indicates FDR corrected p<0.05, Mann-Whitney U test).

#### Overall inflammatory response to TBI is dependent on age

In exploratory analysis, we trained an Elastic Net model on a normative cohort tested on the NULISA™ Inflammation panel to derive age-related coefficients for each predictor marker (*Supplementary Information*). We then applied these coefficients to our data to derive “Inflammation Age”, and “age gap” (“inflammation age” minus calendar age) for each participant. The average “age gap” was significantly higher in the TBI group, compared to CON (t=2.73, p=0.008) but not NTT (t=1.21, p=0.23). The “age gap” was not different between NTT and CON. There was a significant association between “age gap” and calendar age (t=–8.426, p<0.001), with a higher “age gap” in younger participants, although the interaction between group and age on “age gap” was not statistically significant (FIG5). There were no significant correlations between Abbreviated Injury Score (AIS) and Age in the TBI group, nor between between “age gap” and subacute MRI measures, neuronal/astroglial markers or 6-month GOS-E.

**Figure 5:**
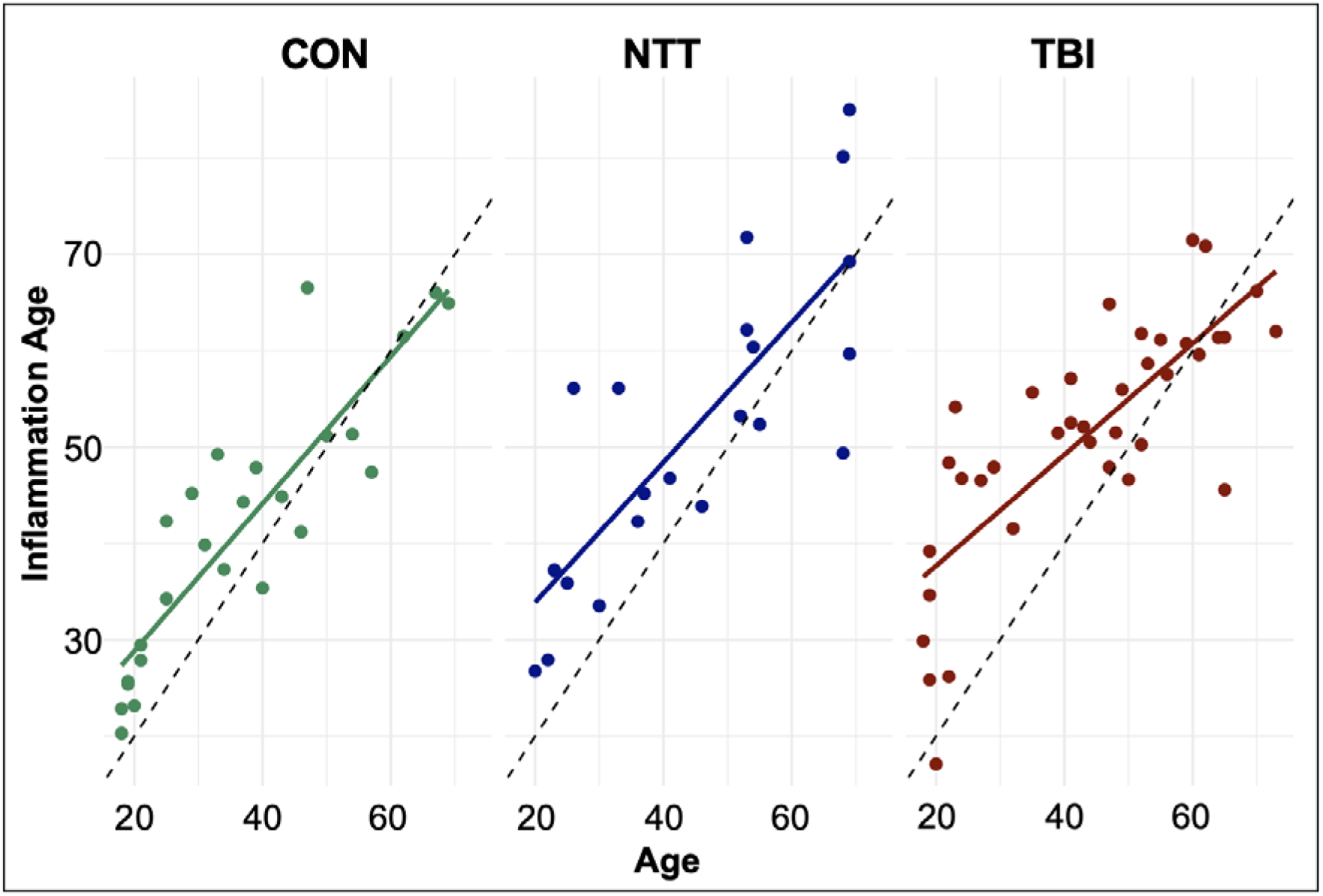
“Inflammation Age” against calendar Age by group. “Inflammation Age” plotted against calendar Age in non-injured controls (CON), non-TBI trauma (NTT) and TBI groups. Each dot represents a participant, with a linear regression line (solid coloured line). The dotted line represents when calendar Age = Inflammation Age.

#### Correlations between proteomic panels and platforms

To assess consistency of protein measurements across different platforms and panels within the same platform, Pearson correlations were calculated for overlapping proteins measured by the Alamar™ NULISA Inflammation and CNS panels, and for the NULISA™ Inflammation and OLINK® Target 96 Inflammation panels (*Supplementary Information*). The majority of overlapping markers on the OLINK® and NULISA™ Inflammation panels demonstrate good correlations, with all but one having r>0.5. The exception was IL18, with r=0.38. All but one overlapping marker on the NULISA™ CNS and Inflammation panels have r>0.7. The exception was CCL22, with r=0.498.

## DISCUSSION

Using the high-dimensional NULISA™ Inflammation proteomic panel for the first time in acute TBI, we demonstrate a marked acute inflammatory response to moderate-severe TBI. Most post-TBI abnormalities in inflammatory markers comprise a general injury response, but several inflammatory markers have TBI-specific derangement. The most robustly TBI-specific inflammatory response is reflected in raised plasma levels of calcium signalling, NF-kB and IL1 pathway markers, as well as GFAP. Proteins involved in the TBI-specific inflammatory response are associated with neuroimaging measures of resulting lesional and white matter injury, whilst the non-specific injury response (IL33) is associated with functional outcome. We also calculated an “inflammation age”, which was increased in acute TBI, with greater increases seen in younger patients. Our results identify specific inflammatory pathways that may drive the acute post-TBI response, and their associations with resulting structural injury and outcomes.

### The TBI-specific acute peripheral inflammatory response includes calcium signalling, NF-kB and IL1 pathways

The novel findings from this study are robust TBI-specific elevation of proteins involved in neuronal calcium signalling (VSNL1), NF-kB inflammatory signalling (IKBKG) and IL1 (IL1RN/IL-1Ra) pathways. In accordance with the wider literature, we also showed a very TBI-specific elevation of plasma GFAP, thought to reflect the extent of intracranial injury^23,24^.

IL1 signalling is considered pro-inflammatory, with IL1b produced by a wide range of inflammatory cells and microglia^25,26^. IL1 mediates a wide range of processes relevant to TBI pathophysiology e.g. reduced BDNF signalling, increased IL-8 and IL-6 production. These processes are associated with worse outcomes^1,2^, development of brain oedema, and recruitment of peripheral immune cells^26^. Indeed, increased acute and subacute levels of plasma IL1b in TBI patients are implicated in worse functional outcomes^2,5^. Further, IL1 signalling blockade with IL-1Ra has shown promise as a potential therapeutic approach in TBI. Pre-clinical studies show that early and sustained IL-1Ra administration results in less cortical volume loss and white matter injury in a mouse model of TBI after polytrauma^27^. An early-stage clinical trial of peripheral anakinra (recombinant IL-1Ra) administration in acute human TBI was both safe and modulated the post-TBI neuroinflammatory response^28,29^. We found higher plasma IL1RN/IL-1Ra levels after TBI. IL1RN encodes IL-1Ra, a competitive antagonist of the IL1 receptor, suggesting that anti-inflammatory signalling is increased after TBI. Given that pro-inflammatory signalling is associated with worse TBI outcomes, our finding that higher plasma IL1RN/IL-1Ra was associated with lower white matter FA, indicating more white matter injury, is unexpected. This may indicate that the plasma IL1RN/IL-1Ra response reflects attempts to counterbalance peripheral pro-inflammatory signalling from other pathways. In keeping with this, IL1RN/IL-1Ra levels were strongly correlated with pro-inflammatory pathway proteins (e.g. IL6, LIF).

A downstream consequence of IL1, and also TNF, signalling is increased NFkB transcription (so-called ‘canonical’ NF-kB activation)^25^. NFkB activation is implicated in a range of chronic human inflammatory diseases^30,31^. We observed TBI-specific increases in plasma IKBKG/NEMO, an essential component of IKK complex-mediated NF-kB activation. This is the first demonstration of peripheral activation of the IKBKG/NEMO pathway in acute human TBI, and complements the pre-clinical literature which reports sustained and widespread activation of NF-kB in the brain after TBI^32^ associated with worse oedema^33^.

Blockade of specific elements of the NF-kB pathway have also been shown to mitigate damage from trauma^34^. On the other hand, NF-kB activation is also implicated in anti-inflammatory processes^31^, and neuronal NF-kB activation may actually be protective in trauma^35^. Therefore, the TBI-specific increase in plasma IKBKG/NEMO may reflect either or both increased pro-inflammatory signalling that is detrimental to clinical outcomes or attempts to mitigate the effects of trauma. Of note, the NF-kB pathway is instrumental in splenic macrophage release of pro-inflammatory cytokines, and rodent model studies show early, and persistent, splenic immune changes after TBI^36,37^, whilst splenectomy immediately post-TBI downregulates NF-kB signalling in the brain. Therefore, this increase in plasma NF-kB may support splenic activation early after human TBI. The broad range of pathways downstream of NF-kB activation indicates that its activation in TBI is unlikely to be a simplistic binary effect. A wide spectrum of pro-inflammatory signals can activate NF-kB, including DAMPs (damage-associated molecular pattern molecules) that are ubiquitous with tissue injury and cell necrosis. Therefore, the TBI-specificity of IKBKG/NEMO activation suggests that the myriad pathways to NF-kB activation are differentially affected by TBI and peripheral injury, which merits further exploration in mechanistic studies.

Plasma levels of the calcium-sensing protein VSNL1 were specifically increased after TBI. Increased plasma levels are also seen in stroke^38^ and neurodegenerative disease^39^. Therefore, elevated VSNL1 levels post-TBI may simply reflect accumulation in plasma due to neuronal death^40^. This would tally with our finding that higher acute VSNL1 levels were positively associated with greater resulting lesion volume. Alternatively, clinical genetic studies suggest that VSNL1 may be linked to molecular mechanisms of Alzheimer’s disease (e.g. increased amyloid precursor protein expression)^41^, and a VSNL1 single-nucleotide polymorphism is associated with cognitive performance in schizophrenia^42^. Disrupted neuronal calcium signalling is recognised after TBI, and is implicated in neuronal death, microglial recruitment and synaptic loss^43,44^. Therefore, elevated peripheral VSNL1 levels may also reflect disrupted calcium signalling that is relevant to the later neuropsychiatric consequences of TBI.

### Derangements in plasma levels of IL33 pathway proteins is associated with functional outcomes

IL33 is an alarmin, proteins that are widely and rapidly released in response to cellular stress or damage^45^. Our finding of increased levels of IL33 and its receptor, IL1RL1, after both acute TBI and NTT is therefore unsurprising. Full-length IL33 is further cleaved to a mature form with high affinity for its membrane-bound receptor (ST2L). This binding results in more Th2 (T-helper Type 2 cell) responses, leading to more anti-inflammatory cytokine release^46^. IL1RL1/soluble ST2 (sST2) is a decoy receptor for IL33, preventing its binding to ST2L, and thus favouring a pro-inflammatory shift in cytokine balance (e.g. increased TNF production)^46^. Indeed, higher blood sST2 levels are associated with a wide range of inflammatory and chronic conditions e.g. disease activity inflammatory bowel disease^47^, and all-cause mortality in heart failure^48^. In this context, our finding of increased peripheral IL1RL1/sST2 after TBI may therefore represent a mechanism to explain why TBI leads to increased risk of cardiovascular disease and all-cause mortality^49–51^.

The NULISA™ Inflammation assay detects both full-length and cleaved IL33, and higher acute levels were seen in TBI patients with POOR GOS-E outcome category than in those with GOOD, in line with previous work showing worse outcomes with higher acute plasma IL33 in a severe TBI cohort^52^. In contrast, another clinical study found that higher IL1RL1/sST2 (i.e. inhibitor of IL33 signalling) was predictive of poor prognosis post-TBI^53^, whilst pre-clinical studies have reported that IL33 signalling seems to be protective post-TBI^54,55^, with increased central and peripheral Treg (regulatory T-cells) numbers and activity a potential mechanism^54^. This is a particularly pertinent finding, given that increasing brain Treg numbers through enhancing brain delivery of IL2 is protective against neuroinflammation in preclinical TBI models^56^. Therefore, it may be that it is the relative, rather than actual, levels of IL33/ST2L and IL33/sST2 activity which is important for determining outcomes. The natural history and mechanistic contributions of this signalling pathway to post-TBI outcomes is an important area for future research, and would benefit from assays that can differentiate between full-length and cleaved IL33 forms. The association of IL33 and IL1RL1/ST2 levels with TBI outcomes also demonstrates that an inflammatory response does not need to be specific to TBI in order to be clinically relevant in TBI.

Our study did not find that plasma GFAP was associated with functional outcome in either univariate or multivariate analyses, which is surprising given the broader literature suggesting that it is^23,57^. This may reflect our relatively small sample size or the fact that we selected this sample based on excluding GFAP outliers in our data. This may have led to restricted variability in GFAP and outcomes within the TBI group, such that a relationship could not be detected.

### The inflammatory response to TBI is co-ordinated and dependent on age

Deranged post-TBI inflammatory markers represent multiple proteins from similar families and pathways (e.g. IL6 and LIF; IL33 and IL1RL1/sST2), and there are also strong positive correlations between markers (e.g. IL6 with IL33, IL1RN). Interventional strategies could thus be targeted to upstream regulatory pathways, such as NfkB, rather than individual proteins. There is a wealth of further work to be done on elucidating the range, extent and natural history of post-TBI inflammatory pathways, and also investigating whether different pathways are affected by different types of injuries, or associated with different clinical outcomes.

In the theory of “inflammaging”, aging can be defined by chronically raised levels of pro-inflammatory markers^58^. We found increased “inflammation age” post-TBI, that is, participants have an older age, based on their inflammation markers, than their calendar age. In the acute context of the present study, this finding likely reflects immediate immune activation, rather than truly ‘aging’. Our previous work assessing ‘brain age’, based on MRI measures of grey and white matter volume, suggested accelerated brain atrophy in chronic TBI survivors^59^. Longitudinal assessment of “inflammation age” would help us understand if TBI also changes trajectory of “inflammaging”. We found that “inflammation age” was age-dependent, with younger TBI participants having a much higher inflammation ‘age gap’ suggestive of overall more derangement in inflammatory marker levels. This implies that while ‘inflammaging’ is characterised by increased pro-inflammatory markers, the dynamic inflammatory response to a major non-infectious insult like TBI may be attenuated in older age. The influence of age on inflammation has been reported previously in neurodegenerative disease e.g. fewer activated microglia are seen in brains of younger patients with Alzheimer’s disease^60^. The pathophysiological and clinical implications of age on TBI-related inflammation, including accounting for potential confounds such as frailty and comorbidity, requires much further exploration.

### Limitations and Future Directions

Our small sample size means that detailed multivariate analyses to investigate the independent associations or prognostic role of specific proteins with specific outcomes were not possible. We cannot control for potential factors e.g. initial injury severity, that may confound some of our findings. The small numbers also meant that we dichotomised the GOS-E, rather than having more categories or using the raw score, which is a very blunt assessment of outcome, particularly as it was not a scaled dichotomisation. Replication with these high-dimensional proteomic platforms is needed in larger cohorts, where such nuanced analyses are possible, to confirm our findings.

The median sampling time was ∼1 day earlier in TBI, compared to NTT, participants. Cytokine and inflammatory protein levels change rapidly, and our adjustments for sample day in differential expression analysis indicated that differences in levels of some proteins (e.g. IL6) between NTT and TBI levels may be more attributable to differences in sampling time than true TBI-specificity. Future studies investigating post-TBI inflammation in the acute setting should take care of timepoint matching. Nevertheless, understanding the non-TBI-specific injury inflammatory response is also important, as our data indicates that this can influence functional outcomes.

It was not possible to explicitly match the groups for comorbidities and types of extracranial injury during recruitment, which are two things that may influence inflammatory response to injury. In larger studies, analyses could adjust for these factors and/or perform sensitivity analyses to interrogate their potential influence on post-TBI inflammation. We are not able to determine, in our study, the provenance of deranged inflammatory markers, nor whether their derangement is causally related to downstream pathology or bystanders. Careful mechanistic work, for example with interventional studies in pre-clinical models, would increase our understanding of this^61^.

Finally, the relative units of the assays in our study limit direct clinical use. Confirmation testing with quantitative assays, for example to determine plasma level thresholds most associated with poor outcomes, would maximise the clinical utility of proteomic work.

A priority for future studies should be investigation of post-TBI inflammation in longitudinal cohorts, as the temporal evolution of post-TBI systemic inflammation is not well understood. Where this has been looked at, raised levels of pro-inflammatory cytokines appear to be raised even several weeks after TBI^2^. On the other hand, it cannot be assumed that all pro-inflammatory processes follow the same trajectory, or that raised systemic cytokine levels reflect detrimental processes at every timepoint. For example, plasma VEGF-A, a multi-functional vascular growth factor, peaks approximately 2 weeks after TBI, and high levels at this timepoint are associated with worse outcomes, but high levels a week later are associated with better outcomes^3,62^. More diverse cohorts (e.g. milder TBIs) are also needed to investigate the broader clinical relevance of our findings outside of the acute, severe setting.

Further, it cannot be assumed that plasma levels are purely reflective of central processes^63^. Where clinical studies have assessed central inflammation, this has been informative about relationships to outcomes that were not identified by, or came to different conclusions to, studies examining peripheral inflammation markers^4,7,12,64^. Measurements of neuroinflammation e.g. measuring inflammatory markers in cerebrospinal or microdialysate (brain interstitial fluid) would also be very useful for future studies, to clarify the cross-talk between systemic and central inflammation post-TBI is unclear.

### Conclusions

We used a high-dimensional proteomic approach to identify proteins involved in the acute inflammatory response to TBI, demonstrating both TBI-specific and non-specific responses. We additionally showed that the inflammatory response is age-dependent, and some of the proteins are associated with resultant white matter injury and functional outcomes. Future work should confirm the findings from this exploratory study, and extend the mechanistic understanding of how derangements in these inflammatory markers are related to patient or injury characteristics and clinical outcomes.

## Data Availability

Research data is available on request to corresponding author (subject to data sharing agreement), and in process of being uploaded to the TBI-REPORTER DPUK Platform (https://tbi-reporter.uk/data-hub/apply-for-data-access/). Original biofluid samples are not available.

## DECLARATIONS

### Ethical Approval and Consent to Participate

Participants provided written informed consent or their professional/personal consultees confirmed participant’s likely willingness to participate. All procedures were approved by the local Research Ethics Committee (IRAS no. 230221), in accordance with the Declaration of Helsinki and UK Human Tissue Authority guidelines.

### Consent for publication

All authors have read the manuscript and consent to its publication.

### Author contributions (CRediT)

Conceptualization: LML, AH, HZ, DJS

Methodology: LML, AH, HZ, NG, KZ, GB, DJS

Validation: EK, AH, HZ, FM, KZ

Data curation: LML, EK, AH, NG, KZ

Formal analysis: CX, LML, ES, KZ, NG

Investigation: LML, EK, AH, NG, KZ, EG, FM, SM, GB, DJS

Resources: LML, AH, HZ, GB, DJS

Writing – original draft: LML, CX

Writing – review and editing: All authors

Supervision: AH, HZ, GB, DJS

Project administration: KZ, NG, GB, DJS

Funding acquisition: GB, DJS

## Acknowledgements

We thank Alamar for providing complimentary testing.

