## Supplementary for "Systemic inflammation and its associations in acute moderate-severe Traumatic Brain Injury: a cross-sectional study"

**Supplementary Information**

**SUPPLEMENTARY METHODS**

**Participants**

Inclusion criteria: adults aged 18-80 years old. TBI patients were included if they fulfilled the Mayo Criteria for moderate-severe TBI (comprising assessment of duration of loss of consciousness, post-traumatic amnesia, Glasgow Coma Scale and initial CT imaging findings). Participants were approached and enrolled only once admitted into hospital, so patients who died from their TBI prior to or very shortly after their arrival into hospital could not practically be included, even though death from TBI fulfils Mayo criteria for moderate-severe TBI.

Exclusion criteria were: pre-existing neurological disease, previous TBI requiring hospitalisation, current drug or alcohol abuse, pregnancy, previous significant disability from any cause. Additional exclusion criteria for the NTT cohort were: any suspicion of TBI (from either mechanism or clinical presentation), and any spinal cord injury

**Full comorbidity list for TBI participants**

| **Patient** | **Comorbidities** |
| --- | --- |
| 1 | Non-insulin dependent Type II Diabetes |
| 2 | Malnutrition |
| 3 | Non-metastatic tumour, Hypertension, Moderate chronic obstructive pulmonary disease |
| 4 | Arrythmia |
| 5 | Hypertension |
| 6 | Hypertension, Non-insulin dependent Type II Diabetes |
| 7 | Arrythmia, Asthma |
| 8 | Previous alcohol/drug misuse, metabolic endocrine disorder |
| 9 | Arrhythmia |
| 10 | Prior alcohol/drug misuse |
| 11 | Prior alcohol/drug misuse |
| 12 | Non-metastatic tumour |

**Sample Selection**

Samples used were from participants from BIO-AX-TBI subset that were included in our previous paper reporting results from testing on the Alamar NULISA™ CNS Diseases panel^1^, with full sample selection details reported therein. 86 participants in total were included in this publication, but 1 CON and 1 TBI participant did not have enough remaining sample to test on Alamar NULISA™ Inflammation panel for this study. This resulted in only samples form 84 participants included in this study.

**Full list of analysed proteins on the Alamar NULISA™ Inflammation Diseases panel**

| AGER | CLEC4A | ICOSLG | IL36G | SDC1 |
| --- | --- | --- | --- | --- |
| AGRP | CNTF | IFNA1 | IL3RA | SELE |
| ANGPT1 | CRP | IFNA1\|IFNA13 | IL4 | SELP |
| ANGPT2 | CSF1 | IFNA2 | IL4R | SIRPA |
| ANXA1 | CSF1R | IFNB1 | IL5 | SLAMF1 |
| AREG | CSF2 | IFNG | IL5RA | SPP1 |
| BDNF | CSF2RB | IFNL1 | IL6 | TAFA5 |
| BMP7 | CSF3 | IFNL2\|IFNL3 | IL6R | TEK |
| BST2 | CSF3R | IFNW1 | IL6ST | TGFB1 |
| C1QA | CST7 | IKBKG | IL7 | THBS2 |
| CALCA (*procalcitonin)* | CTF1 | IL10 | IL7R | THPO |
| CCL1 | CTLA4 | IL10RB | IL9 | TIMP1 |
| CCL11 | CTSS | IL11 | IRAK4 | TIMP2 |
| CCL13 | CX3CL1 | IL12B | KDR | TLR3 |
| CCL14 | CXADR | IL12RB1 | KITLG | TNF |
| CCL15 | CXCL1 | IL12p70 | KLRK1 | TNFRSF11A |
| CCL16 | CXCL10 | IL13 | KNG1 | TNFRSF11B |
| CCL17 | CXCL11 | IL13RA2 | LAG3 | TNFRSF13B |
| CCL19 | CXCL12 | IL15 | LAMP3 | TNFRSF13C |
| CCL2 | CXCL13 | IL15RA | LCN2 | TNFRSF14 |
| CCL20 | CXCL14 | IL16 | LGALS9 | TNFRSF17 |
| CCL21 | CXCL16 | IL17A | LIF | TNFRSF18 |
| CCL22 | CXCL2 | IL17A\|IL17F | LILRB2 | TNFRSF1A |
| CCL23 | CXCL3 | IL17B | LTA | TNFRSF1B |
| CCL24 | CXCL5 | IL17C | LTA\|LTB | TNFRSF21 |
| CCL25 | CXCL6 | IL17F | MERTK | TNFRSF4 |
| CCL26 | CXCL8 | IL17RA | MICA | TNFRSF8 |
| CCL27 | CXCL9 | IL17RB | MICB | TNFRSF9 |
| CCL28 | EGF | IL18 | MIF | TNFSF10 |
| CCL3 | EPO | IL18BP | MMP1 | TNFSF11 |
| CCL4 | FASLG | IL18R1 | MMP12 | TNFSF12 |
| CCL5 | FGF19 | IL19 | MMP3 | TNFSF13 |
| CCL7 | FGF2 | IL1B | MMP8 | TNFSF14 |
| CCL8 | FGF21 | IL1R1 | MMP9 | TNFSF15 |
| CD200 | FGF23 | IL1R2 | MPO | TNFSF18 |
| CD200R1 | FLT1 | IL1RL1 | MUC16 | TNFSF4 |
| CD27 | FLT3LG | IL1RN *(IL-1Ra)* | NAMPT | TNFSF8 |
| CD274 | FLT4 | IL2 | NCR1 | TNFSF9 |
| CD276 | FTH1 | IL20 | NGF | TREM1 |
| CD3E | FURIN | IL22 | NTF3 | TREM2 |
| CD4 | GDF15 | IL23 | OSM | VCAM1 |
| CD40 | GDF2 | IL24 | OSMR | VEGFA |
| CD40LG | GFAP | IL27 | PDCD1 | VEGFC |
| CD46 | GRN | IL2RA | PDCD1LG2 | VEGFD |
| CD70 | GZMA | IL2RB | PDGFA | VSNL1 |
| CD80 | GZMB | IL32 | PDGFB | VSTM1 |
| CD83 | HAVCR1 | IL33 | PTX3 | WNT16 |
| CD93 | HGF | IL34 | S100A12 | WNT7A |
| CEACAM5 | HLA-DRA | IL36A | S100A9 | mCherry |
| CHI3L1 | ICAM1 | IL36B | SCG2 |  |

NB: protein names as termed by company brochure information (<https://alamarbio.com/products-and-services/nulisa-inflammation-panel/>). Some relevant annotations provided in italics within brackets.

**MRI Lesion analysis**

We calculated lesion volume from manually drawn masks, using T1w and T2 FLAIR scans, and volumes extracted from the masks with *fslstats* from the FSL imaging analysis software package^24^. KZ, FM and NG carried out manual lesion segmentation, initially working together on a small subset to ensure consistency of approach, before working independently. NG is a neurologist, and all are experienced neuroimaging researchers.

**Inflammation Age Model training**

We used Elastic net (*cv.glmnet* function, alpha=0.1, nfold=50) to deal with multicollinearity within proteomic data. Iteration over alpha values identified 0 (i.e. Ridge regression) as the optimal alpha, but this does not allow feature selection so included many proteins with minimal association with age in the resulting model, hence we chose to use Elastic Net with an alpha value close to 0. Before training, we removed data pertaining to 4 proteins that did not appear in one of our dataset or the training dataset (mCherry, PGF, TGFB3, TLSP). We also removed GFAP for the core model, as this is a very strongly TBI-specific intracranial injury marker, to ensure that any differences in “Inflammation Age” is not purely driven by GFAP.

**SUPPLEMENTARY RESULTS**

**Differential Expression coefficients**

Markers from the Alamar NULISA™ Inflammation panel showing statistically significantly different plasma levels between the CON, NTT and TBI groups in the core differential expression analysis, where the coefficient for at least one pairwise comparison is ≥1 (i.e. at least a 2-fold change in plasma levels).

| **Marker** | **TBI_vs_CON**  **coefficient** | **TBI_vs_NTT**  **coefficient** | **NTT_vs_CON**  **coefficient** | **F** | **adj.P.Val** |
| --- | --- | --- | --- | --- | --- |
| GFAP | 6.959031958 | 6.54848196 | 0.410549998 | 536.7725 | 4.35E-45 |
| IL6 | 5.621254093 | 1.714201808 | 3.907052284 | 80.00377 | 8.12E-18 |
| TNFSF11 | -2.636285234 | -0.866249212 | -1.770036022 | 73.85161 | 4.52E-17 |
| CRP | 4.892048714 | 0.338050757 | 4.553997958 | 52.76211 | 1.31E-13 |
| LIF | 3.443338753 | 1.211568166 | 2.231770587 | 49.21303 | 5.05E-13 |
| PTX3 | 2.768968141 | 1.230972191 | 1.53799595 | 46.64105 | 1.37E-12 |
| IL1RL1 | 3.033082506 | 1.842187903 | 1.190894602 | 41.55395 | 1.34E-11 |
| IL33 | 2.386805143 | 1.157050362 | 1.229754781 | 39.62023 | 3.09E-11 |
| TNFSF14 | 2.494147085 | -0.046485731 | 2.540632816 | 37.18253 | 9.59E-11 |
| CCL7 | 2.222586024 | 0.67436173 | 1.548224294 | 34.56813 | 3.45E-10 |
| TAFA5 | -1.48769771 | -0.819532139 | -0.668165571 | 33.01609 | 7.32E-10 |
| IL1RN | 2.403091444 | 1.415441138 | 0.987650306 | 25.37284 | 5.73E-08 |
| S100A12 | 1.707652267 | -0.930019086 | 2.637671353 | 24.49751 | 8.57E-08 |
| CSF3 | 1.978051635 | 0.970213575 | 1.00783806 | 24.47433 | 8.57E-08 |
| CCL23 | 1.196019466 | 0.520461556 | 0.67555791 | 24.31693 | 8.82E-08 |
| IL15 | 0.876276431 | 0.262267886 | 0.614008545 | 24.17749 | 8.98E-08 |
| CHI3L1 | 2.932244216 | 1.430818622 | 1.501425593 | 24.07348 | 9.07E-08 |
| VSNL1 | 1.614388867 | 1.221776099 | 0.392612768 | 23.1352 | 1.55E-07 |
| TNFSF10 | -1.359350017 | -0.565445822 | -0.793904195 | 21.93531 | 3.17E-07 |
| OSM | 1.678650855 | 0.81283817 | 0.865812686 | 19.83083 | 1.21E-06 |
| HGF | 1.154849343 | 0.472205187 | 0.682644156 | 18.47327 | 2.53E-06 |
| WNT16 | -1.242193545 | -0.544227606 | -0.697965939 | 17.60627 | 4.26E-06 |
| FURIN | 1.097379948 | 0.249445866 | 0.847934082 | 16.35803 | 8.9E-06 |
| AREG | 1.017713021 | 0.502881854 | 0.514831167 | 15.78576 | 1.3E-05 |
| IL10 | 1.87892066 | 0.776101362 | 1.102819298 | 15.26247 | 1.73E-05 |
| SPP1 | 1.175912976 | 0.743759354 | 0.432153622 | 15.08395 | 1.91E-05 |
| CALCA | 1.716426726 | 1.283211158 | 0.433215568 | 14.02397 | 3.84E-05 |
| IL18BP | 1.030354474 | 0.475043489 | 0.555310985 | 13.07604 | 7.07E-05 |
| CST7 | 1.350765398 | 0.592117016 | 0.758648383 | 12.63166 | 8.89E-05 |
| EPO | 1.639007707 | 0.709304784 | 0.929702922 | 12.25219 | 0.000116 |
| MMP8 | 1.482193799 | 0.944035503 | 0.538158296 | 11.57822 | 0.000187 |
| BST2 | 1.286479709 | 0.545588334 | 0.740891375 | 10.20593 | 0.000511 |
| IL4 | -1.029220317 | -0.318248839 | -0.710971478 | 8.697466 | 0.001591 |
| IKBKG | 1.028579444 | 1.51457518 | -0.485995736 | 8.669876 | 0.0016 |
| TIMP1 | 1.002000212 | 1.050735608 | -0.048735395 | 8.529702 | 0.001685 |
| FGF21 | 1.367915594 | 0.92521323 | 0.442702364 | 6.64181 | 0.006694 |
| IL18 | 0.327181385 | 1.035384245 | -0.70820286 | 6.470218 | 0.007662 |
| MIF | 0.044954302 | 1.522164374 | -1.477210072 | 5.881164 | 0.012166 |
| IRAK4 | 0.318686157 | 1.809664169 | -1.490978012 | 4.188753 | 0.043044 |

**Boxplots showing plasma levels of proteins with TBI specificity (from core differential expression analysis)**

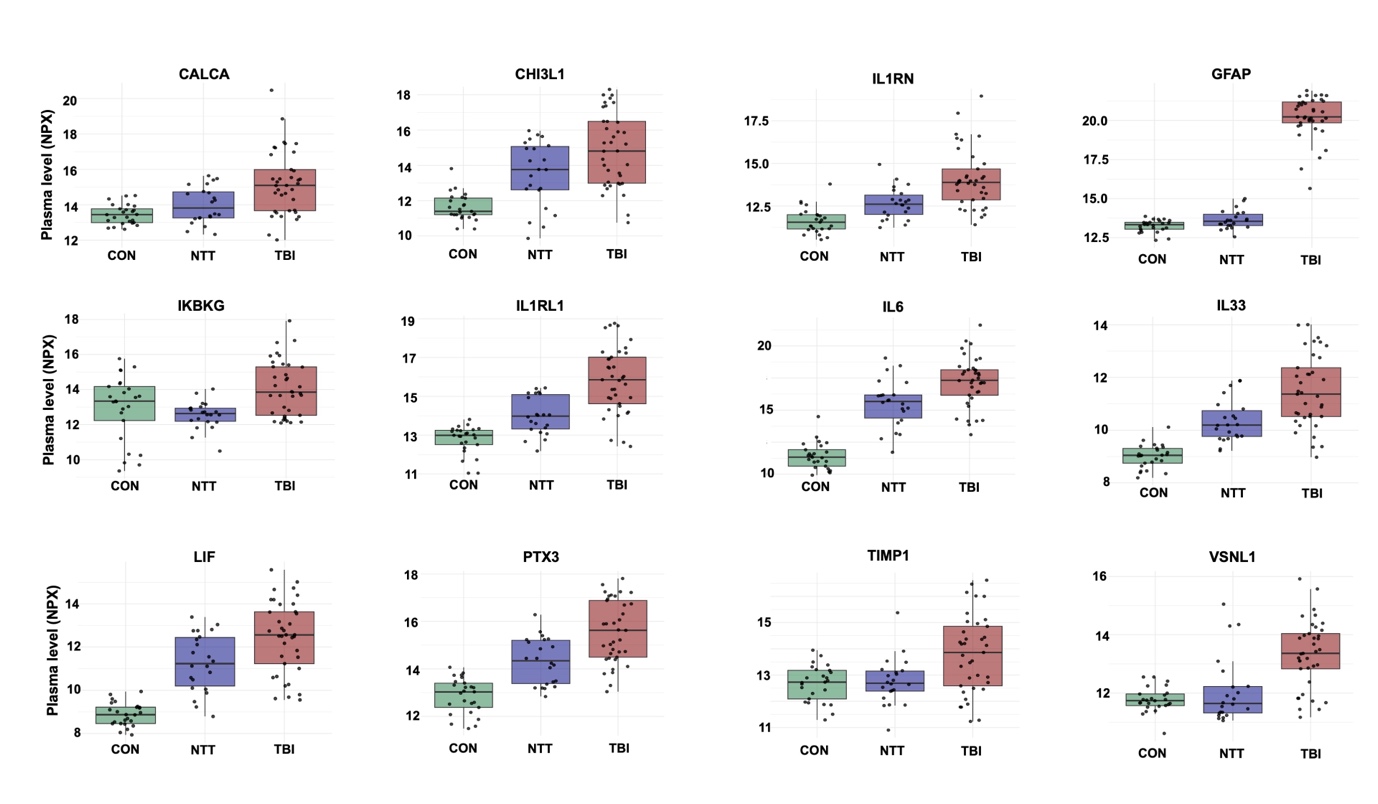

*Figure 1: Plasma protein levels in CON, NTT and TBI groups for proteins identified by the core differential expression analysis to show TBI specificity in derangement in  acute post-TBI plasma levels. The y-axis units are NULISA protein quantification (NPQ) units. Each dot represents a participant, box limits are the interquartile range (IQR), whisker limits are 1.5x IQR, midline is median.*

**Coefficient values for each protein derived from the Elastic Net model for “Inflammation Age”**

| **Coefficient** | **Marker** |  | **Coefficient** | **Marker** |
| --- | --- | --- | --- | --- |
| 0.148468 | ANGPT1 |  | -0.83955 | AGER |
| 0.746315 | C1QA |  | -2.89134 | AGRP |
| 0.563561 | CCL1 |  | -0.27227 | ANXA1 |
| 3.621265 | CCL11 |  | -0.30029 | AREG |
| 0.450953 | CCL13 |  | -3.12124 | BMP7 |
| 0.774347 | CCL14 |  | -2.13563 | BST2 |
| 0.427932 | CCL15 |  | -2.72372 | CCL21 |
| 1.118368 | CCL27 |  | -1.2807 | CCL22 |
| 0.557226 | CCL28 |  | -1.38446 | CCL25 |
| 0.068128 | CCL3 |  | -0.46247 | CCL26 |
| 0.12117 | CCL4 |  | -0.24301 | CCL5 |
| 0.483446 | CD276 |  | -0.4365 | CD200R1 |
| 0.14879 | CD70 |  | -1.76368 | CD274 |
| 0.559949 | CD80 |  | -0.06802 | CD3E |
| 0.94767 | CEACAM5 |  | -0.46276 | CD4 |
| 1.473814 | CHI3L1 |  | -1.24381 | CD83 |
| 0.013521 | CTLA4 |  | -0.46543 | CLEC4A |
| 0.94707 | CXCL10 |  | -0.15692 | CSF1R |
| 0.085394 | CXCL16 |  | -0.34902 | CSF2RB |
| 1.011506 | CXCL9 |  | -0.5717 | CSF3 |
| 0.606847 | FGF19 |  | -0.70311 | CST7 |
| 0.182349 | FGF2 |  | -3.46289 | CX3CL1 |
| 1.00073 | FGF21 |  | -0.03851 | CXADR |
| 0.959784 | FLT3LG |  | -0.11812 | CXCL11 |
| 0.531392 | FTH1 |  | -0.16839 | CXCL12 |
| 1.474009 | FURIN |  | -0.48121 | CXCL13 |
| 0.028768 | GDF15 |  | -0.42313 | CXCL14 |
| 0.522604 | GDF2 |  | -0.11396 | CXCL2 |
| 0.33754 | GRN |  | -0.44407 | CXCL6 |
| 0.406032 | HAVCR1 |  | -0.19405 | CXCL8 |
| 1.579389 | HLA-DRA |  | -0.54857 | FASLG |
| 0.300606 | IFNW1 |  | -0.7223 | FGF23 |
| 0.493948 | IKBKG |  | -1.06363 | GZMA |
| 0.803346 | IL12p70 |  | -1.41083 | ICOSLG |
| 0.740556 | IL12RB1 |  | -0.78182 | IFNG |
| 1.663092 | IL15RA |  | -0.03109 | IL11 |
| 1.348668 | IL16 |  | -0.98997 | IL15 |
| 0.079594 | IL18 |  | -0.1711 | IL17A |
| 0.870534 | IL1R1 |  | -0.69948 | IL17B |
| 2.111994 | IL2 |  | -0.07103 | IL17C |
| 0.453203 | IL3RA |  | -0.11993 | IL17F |
| 0.707202 | IL5 |  | -1.1494 | IL17RA |
| 1.436049 | IL5RA |  | -0.97204 | IL18R1 |
| 0.554497 | IL9 |  | -1.89915 | IL1R2 |
| 0.272362 | IRAK4 |  | -1.10397 | IL1RL1 |
| 0.698464 | KDR |  | -0.77164 | IL1RN |
| 1.801685 | KITLG |  | -0.20226 | IL22 |
| 0.8806 | LAG3 |  | -0.33597 | IL24 |
| 1.505282 | LAMP3 |  | -0.29567 | IL27 |
| 0.290894 | LCN2 |  | -0.12945 | IL32 |
| 1.063657 | LGALS9 |  | -0.25121 | IL33 |
| 0.839428 | LILRB2 |  | -0.12325 | IL34 |
| 0.036795 | MICA |  | -0.40921 | IL36B |
| 0.440983 | MMP1 |  | -0.18264 | IL36G |
| 0.639106 | MMP12 |  | -2.21219 | IL4R |
| 0.370126 | MMP9 |  | -0.03149 | IL6R |
| 0.116977 | MPO |  | -1.19994 | IL7R |
| 0.061872 | NAMPT |  | -3.17072 | KLRK1 |
| 0.088783 | NCR1 |  | -0.15677 | KNG1 |
| 2.567337 | OSMR |  | -0.32748 | LTA |
| 1.233216 | SELP |  | -0.03498 | LTA\|LTB |
| 0.053732 | SIRPA |  | -1.10931 | MUC16 |
| 0.314482 | TGFB1 |  | -0.54444 | NGF |
| 0.473034 | TIMP1 |  | -0.61449 | PDCD1 |
| 0.120647 | TNFRSF11B |  | -0.48947 | PDGFA |
| 1.128469 | TNFRSF13B |  | -0.37844 | S100A12 |
| 1.020193 | TNFRSF17 |  | -0.14286 | S100A9 |
| 0.27845 | TNFRSF4 |  | -0.21594 | SCG2 |
| 1.052972 | TNFSF12 |  | -1.69852 | SDC1 |
| 3.51125 | TNFSF15 |  | -1.54206 | SELE |
| 0.262523 | TNFSF4 |  | -1.18966 | TAFA5 |
| 1.267492 | TNFSF9 |  | -0.9249 | THBS2 |
| 1.225644 | TREM1 |  | -1.72751 | THPO |
| 2.901125 | TREM2 |  | -0.0895 | TNF |
| 3.338782 | VCAM1 |  | -1.03536 | TNFRSF13C |
| 4.075214 | VEGFD |  | -0.41355 | TNFRSF1B |
| 1.122709 | WNT16 |  | -1.75824 | TNFRSF8 |
| 0.19267 | WNT7A |  | -1.42601 | TNFSF10 |
|  |  |  | -0.99437 | TNFSF11 |
|  |  |  | -0.15441 | VSNL1 |

**Cross-panel and Cross-platform correlations between overlapping markers assayed on different panels and platforms**

Pearson’s correlation was used.

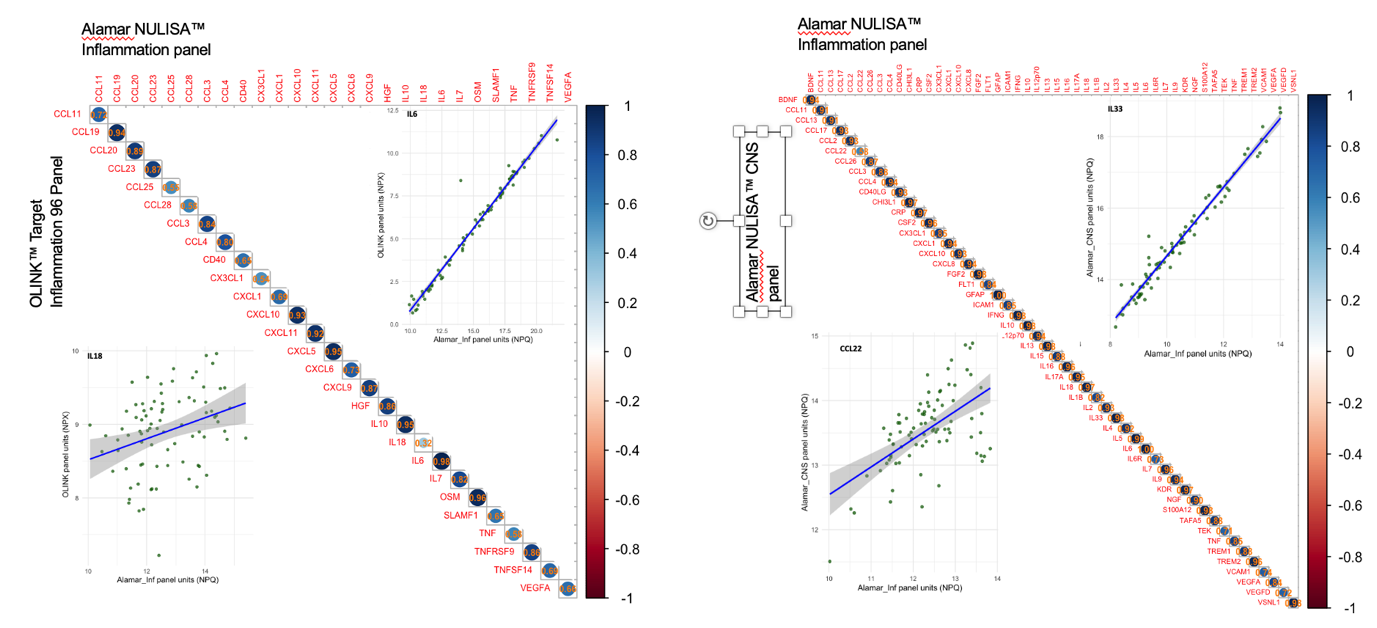

***Figure 2.*** *Pairwise Pearson correlations between protein levels measured by the Alamar™ NULISA Inflammation panel (horizontal axis) and two alternative assays.* ***(A)*** *the OLINK® Target 96 Inflammation panel, and* ***(B)*** *the Alamar™ NULISA CNS Diseases panel. The darker the colour of the circle, the stronger the correlation. Insets are scatterplots for the markers with lowest and highest r values among overlapping proteins. Solid lines represent linear regression fits with 95% confidence intervals (shaded areas).*
